# Caution on Kidney Dysfunctions of COVID-19 Patients

**DOI:** 10.1101/2020.02.08.20021212

**Authors:** Zhen Li, Ming Wu, Jiwei Yao, Jie Guo, Xiang Liao, Siji Song, Jiali Li, Guangjie Duan, Yuanxiu Zhou, Xiaojun Wu, Zhansong Zhou, Taojiao Wang, Ming Hu, Xianxiang Chen, Yu Fu, Chong Lei, Hailong Dong, Chuou Xu, Yahua Hu, Min Han, Yi Zhou, Hongbo Jia, Xiaowei Chen, Junan Yan

## Abstract

**Background:** To date, large amounts of epidemiological and case study data have been available for the Coronavirus Disease 2019 (COVID-19), which suggested that the mortality was related to not just respiratory complications. Here, we specifically analyzed kidney functions in COVID-19 patients and their relations to mortality.

**Method:** In this multi-centered, retrospective, observational study, we included 193 adult patients with laboratory-confirmed COVID-19 from 2 hospitals in Wuhan, 1 hospital in Huangshi (Hubei province, 83 km from Wuhan) and 1 hospital in Chongqing (754 km from Wuhan). Demographic data, symptoms, laboratory values, comorbidities, treatments, and clinical outcomes were all collected, including data regarding to kidney functions. Data were compared among three groups: non-severe COVID-19 patients (128), severe COVID-19 patients (65) and a control group of other pneumonia (28). For the data from computed tomographic (CT) scans, we also included a control group of healthy subjects (110 cases, without abnormalities in the lung and without kidney diseases). The primary outcome was a common presence of kidney dysfunctions in COVID-19 patients and the occurrence of acute kidney injury (AKI) in a fraction of COVID-19 patients. Secondary outcomes included a survival analysis of COVID-19 patients in conditions of AKI or comorbid chronic illnesses.

**Findings:** We included 193 COVID-19 patients (128 non-severe, 65 severe (including 32 non-survivors), between January 6^th^ and February 21^th^,2020; the final date of follow-up was March 4^th^, 2020) and 28 patients of other pneumonia (15 of viral pneumonia, 13 of mycoplasma pneumonia) before the COVID-19 outbreak. On hospital admission, a remarkable fraction of patients had signs of kidney dysfunctions, including 59% with proteinuria, 44% with hematuria, 14% with increased levels of blood urea nitrogen, and 10% with increased levels of serum creatinine, although mild but worse than that in cases with other pneumonia. While these kidney dysfunctions might not be readily diagnosed as AKI at admission, over the progress during hospitalization they could be gradually worsened and diagnosed as AKI. A univariate Cox regression analysis showed that proteinuria, hematuria, and elevated levels of blood urea nitrogen, serum creatinine, uric acid as well as D-dimer were significantly associated with the death of COVID-19 patients respectively. Importantly, the Cox regression analysis also suggested that COVID-19 patients that developed AKI had a ∼5.3-times mortality risk of those without AKI, much higher than that of comorbid chronic illnesses (∼1.5 times risk of those without comorbid chronic illnesses).

**Interpretation:** To prevent fatality in such conditions, we suggested a high degree of caution in monitoring the kidney functions of severe COVID-19 patients regardless of the past disease history. In addition, upon day-by-day monitoring, clinicians should consider any potential interventions to protect kidney functions at the early stage of the disease and renal replacement therapies in severely ill patients, particularly for those with strong inflammatory reactions or a cytokine storm.

**Funding:** None.

## Introduction

By March 11, 2020, the World Health Organization (WHO) has declared that the outbreak of the new coronavirus is a global pandemic. The pathogen of COVID-19, severe acute respiratory syndrome coronavirus 2 (SARS-CoV-2), is highly transmissive^1^ even by asymptomatic infected subjects^2^ and thus very difficult to be contained. Given that the development and trials of drugs and vaccinations still require months or years to be completed and yet cannot be readily applied in all countries, a plausible global public health strategy would be to apply already-established means of interventions to help those severely ill COVID-19 patients by reducing the risk of dying. To do so, specific investigations on possible high-risk mortality factors should be first undertaken. Large-scale epidemiological data^3^ have demonstrated that the mortality rate of SARS-CoV-2 infection is significantly higher for patients with higher ages and/or with comorbid chronic illnesses such as cardiovascular diseases, diabetes, chronic respiratory diseases, hypertension and cancer^4^. However, these basic chronic illnesses are mortality risk factors for many other pathogen infections, commonly exist in old people and are hardly curable. Thus, they obscure the investigation of specific mortality risk factors for COVID-19 patients and made it difficult to justify the rationale and fairness of allocating public medical resources for condition-specific interventions. To address this issue, a recent key report^5^ suggested a number of potential risk factors, including older age, high sequential organ failure assessment score and D-dimer greater than 1 μg/L for identifying COVID-19 patients with poor prognosis. Equally importantly, here our retrospective investigation of COVID-19 patients revealed a specific mortality risk factor: kidney dysfunctions, which generally appeared at first as mild abnormalities and could later develop as clinically-diagnosed acute kidney injury (AKI) in a significant fraction of severely ill patients. The development of AKI in COVID-19 patients is a critical negative prognostic factor for survival, which, unlike other known negative prognostic factors, is possibly curable by interventions.

## Research in context

### Evidence before this study

SARS-CoV-2 uses ACE2 (angiotensin-converting enzyme II) as a cell entry receptor^6^, a cellular mechanism identical to that of the previously known SARS-CoV^7^. However, ACE2 was not exclusively expressed in the respiratory organs of humans. A PubMed database (https://www.ncbi.nlm.nih.gov/gene/?term=59272) shows that ACE2 RNA expressions in gastrointestinal organs (small intestine, duodenum) and urinary organs (kidney) are much higher (nearly 100-fold) than that in respiratory organs (lung). On the other hand of clinical case studies, a detailed retrospective study of the 2003 SARS-CoV outbreak^8^ shows that acute kidney injury was uncommon in SARS patients but carried a formidably high mortality (91.7%, 33 of 36 cases). These lines of evidence prompted us to investigate kidney dysfunctions in COVID-19 patients.

### Added value of this study

This is the first systematic investigation on kidney functions of COVID-19 patients. In many other studies on COVID-19 patients, the kidney functions of patients and the existence of AKI were reported in standard examinations and epidemiological data but without further comparative analysis. Here, we involved a control group of patients of other commonly known pneumonia and found that COVID-19 patients exhibited more significant kidney dysfunctions. Our study features a live tracking of kidney functions in addition to a snapshot at hospital admission. Furthermore, we managed to collect kidney CT scan data in the same datasets which were not shown in previous reports.

### Implications of all the available evidence

Although pneumonia is the most prevalent characteristics of COVID-19, our study, from a different point of view, revealed kidney dysfunctions in COVID-19 patients that were generally mild but significantly worse than that in patients of other pneumonia. A fraction of COVID-19 patients developed AKI, which, in a survival analysis, showed much more critical prognosis of morality than that of comorbid chronic illnesses that were already known to relate to a high mortality rate. These results together suggest exercising a high degree of caution in monitoring the kidney functions of COVID-19 patients, and applying potential interventions to protect kidney functions at the early stage of the disease and, if possible, renal replacement therapies (RRT) in severely ill patients, particularly for those with strong inflammatory reactions or a cytokine storm.

## Methods

### Study design and participants

This is a retrospective study performed at four hospitals in Hubei province and in Chongqing city. A total number of 193 patients with confirmed COVID-19 pneumonia admitted to the Wuhan Tongji hospital, Wuhan Pulmonary Hospital, Huangshi Central Hospital and Chongqing Southwest hospital between January 6^th^ and February 21^th^,2020, were enrolled. We also enrolled 28 patients diagnosed of other types of pneumonia (15 patients of viral pneumonia and 13 patients of mycoplasma pneumonia) before the COVID-19 outbreak from Wuhan Tongji Hospital as a control group. The clinical outcomes (namely remained in hospital, discharges, death) were monitored until March 4^th^, 2020. This study has been approved by the Ethics Commissions of Wuhan Tongji Hospital (TJ-IRB20200216). Written informed consent was waived by the Ethics Commission of the designated hospital for emerging infectious diseases. All the authors reviewed the manuscript and vouch for the accuracy and completeness of the data and for the adherence of the study to the protocol.

All COVID-19 patients enrolled in the study were diagnosed according to the latest guidance provided by National Health Commission of the People’s Republic of China (NHC China). The diagnoses were made based on two laboratory investigations: (1) respiratory specimens, including sputum, nasal and pharyngeal swabs, were tested using a COVID-19 nucleic acid detection kit; (2) multiple patchy shadows or ground-glass shadows were detected using lung computed tomographic (CT) scans.

### Procedures

We extracted epidemiological characteristics, demographic characteristics, clinical characteristics, laboratory data, radiological characteristics, treatment and outcomes data from electronic medical records. We recorded the information on age, sex, recent exposure history, underlying comorbidities, clinical symptoms from the onset to hospital admission, physical examination, CT scans, laboratory findings on admission (a complete blood count, blood chemical analysis, coagulation testing, assessment of liver and renal function, and measures of hypersensitive C-reactive protein, procalcitonin, routine urine), treatment measures (antiviral therapy, oxygen therapy, glucocorticoid therapy, respiratory support, renal replacement therapy) and living status (Tables 1, 2 and 3). All the data were checked by a team of trained physicians.

**Table 1:** Demographics, baseline characteristics, clinical characteristics of COVID-19 patients

|  | No. (%) |  |  |  |  |
| --- | --- | --- | --- | --- | --- |
|  | All COVID-19 pneumonia patients (n=193) | Non-severe (n=128) | Severe (n=65) | Patients with other pneumonia (n=28) | P value |
| **Characteristics** |  |  |  |  | CP group vs OP group ; Non-severe vs severe |
| Age, median (IQR), y | 57 (46-67) | 55 (43-66) | 66 (56-71) | 36 (30-54) | P<0.001; P<0.001 |
| Sex |  |  |  |  | P>0.05; P<0.05 |
| Male | 95 (49%) | 56 (44%) | 39 (60%) | 17 (61%) |  |
| Female | 98 (51%) | 72 (56%) | 26 (40%) | 11 (39%) |  |
| Exposure |  |  |  |  |  |
| Local residents of Wuhan | 116 (60%) | 94 (73%) | 22 (34%) |  |  |
| Recently been to Wuhan/Hubei province | 8 (4%) | 3 (2%) | 5 (8%) |  |  |
| Exposure to patients | 31 (16%) | 26 (20%) | 5 (8%) |  |  |
| Medical staff, % | 16 (8%) | 14 (11%) | 2 (3%) | 0 (0%) | P>0.05; P>0.05 |
| **Comorbidities** | 105 (54%) | 62 (48%) | 43 (66%) |  | P>0.05; P<0.05 |
| Nervous system diseases | 5 (3%) | 2 (2%) | 3 (5%) | 0 (0%) | P>0.05; P>0.05 |
| Cardiovascular and cerebrovascular diseases | 70 (36%) | 41 (32%) | 29 (45%) | 4 (14%) | P<0.05; P>0.05 |
| Respiratory system diseases | 27 (14%) | 10 (8%) | 17 (26%) | 6 (21%) | P>0.05; P<0.01 |
| Digestive system diseases | 19 (10%) | 9 (7%) | 10 (15%) | 6 (21%) | P>0.05; P>0.05 |
| Urinary system diseases | 10 (5%) | 4 (3%) | 6 (9%) | 3 (11%) | P>0.05; P>0.05 |
| Reproductive system diseases | 3 (2%) | 2 (2%) | 1 (2%) | 0 (0%) | P>0.05; P>0.05 |
| Endocrine system diseases | 38 (20%) | 24 (19%) | 14 (22%) | 1 (4%) | P>0.05; P>0.05 |
| **Signs and symptoms at admission** |  |  |  |  |  |
| Fever | 171 (89%) | 116 (91%) | 55 (85%) | 18 (64%) | P<0.01; P>0.05 |
| Cough | 134 (69%) | 83 (65%) | 51 (78%) | 22 (79%) | P>0.05; P>0.05 |
| Expectoration | 59 (31%) | 38 (30%) | 21 (32%) | 17 (61%) | P<0.01; P>0.05 |
| Rhinorrhoea | 5 (3%) | 5 (4%) | 0 (0%) | 1 (4%) | P>0.05; P>0.05 |
| Sore throat | 13 (7%) | 7 (5%) | 6 (9%) | 0 (0%) | P>0.05; P>0.05 |
| Dyspnea | 70 (36%) | 30 (23%) | 40 (62%) | 1 (4%) | P<0.01; P<0.001 |
| Chest pain | 13 (7%) | 5 (4%) | 8 (12%) | 3 (11%) | P>0.05; P>0.05 |
| Myalgia | 26 (14%) | 20 (16%) | 6 (9%) | 3 (11%) | P>0.05; P>0.05 |
| Abdominal pain | 8 (4%) | 4 (3%) | 4 (6%) | 0 (0%) | P>0.05; P>0.05 |
| Low back pain | 1 (1%) | 1 (1%) | 0 (0%) | 1 (4%) | P>0.05; P>0.05 |
| Headache | 20 (10%) | 16 (13%) | 4 (6%) | 2 (7%) | P>0.05; P>0.05 |
| Fatigue | 75 (39%) | 44 (34%) | 31 (48%) | 5 (18%) | P>0.05; P>0.05 |
| Dizziness | 13 (7%) | 10 (8%) | 3 (5%) | 2 (7%) | P>0.05; P>0.05 |
| Anorexia | 62 (32%) | 26 (20%) | 36 (55%) | 2 (7%) | P<0.05; P<0.001 |
| Diarrhoea | 35 (18%) | 28 (22%) | 7 (11%) | 1 (4%) | P>0.05; P>0.05 |
| Nausea | 11 (6%) | 5 (4%) | 6 (9%) | 0 (0%) | P>0.05; P>0.05 |
| Vomiting | 5 (3%) | 4 (3%) | 1 (2%) | 0 (0%) | P>0.05; P>0.05 |
| Maximum temperature, °C | 38.5 (37.8-39) | 38.4 (37.7-39) | 38.5 (37.8-39) | 38.5 (36.6-39) | P>0.05; P>0.05 |
| Respiratory rate, median (IQR), breaths per min | 20 (20-24) | 20 (20-22) | 22 (20-28) | 20 (20-20) | P<0.01; P<0.01 |
| Systolic pressure, median (IQR), mm Hg | 124 (116-135) | 122 (116-133) | 129 (120-140) | 112 (105-129) | P<0.01; P>0.05 |
| Diastolic pressure, median (IQR), mm Hg | 77 (70-85) | 77 (71-85) | 75 (70-82) | 76 (68-84) | P>0.05; P>0.05 |
| Heart rate, median (IQR), bpm | 89 (80-99) | 89 (80-98) | 86 (80-99) | 88 (78-107) | P>0.05; P>0.05 |
| **Chest CT findings** |  |  |  |  |  |
| Pneumonia | 193 (100%) | 128 (100%) | 65 (100%) | 28 (100%) | P>0.05; P>0.05 |
| Multiple mottling and ground-glass opacity | 152 (79%) | 115 (90%) | 37 (58%) | 8 (29%) | P<0.001; P<0.001 |

Data are mean (SD), median (IQR) and n (%). CP group=COVID-19 pneumonia group. OP group=other pneumonia group.

**Table 2:** Complications, treatments and clinical outcomes of patients with COVID-19 pneumonia

|  | No. (%)<br>All COVID-19<br>pneumonia<br>patients (n=193) | Non-severe<br>(n=128) | Severe (n=65) | Patients with other<br>pneumonia (n=28) | P value |
| --- | --- | --- | --- | --- | --- |
| **Complications** |  |  |  |  | CP group vs OP<br>group ; Non-severe<br>vs severe |
| ARDS | 54 (28%) | 2 (2%) | 52 (80%) | 1 (4%) | P<0.05; P<0.001 |
| Acute cardiac injury | 24 (12%) | 5 (4%) | 19 (29%) | 2 (7%) | P>0.05; P<0.001 |
| Septic shock | 35 (18%) | 0 (0%) | 35 (54%) | 1 (4%) | P>0.05; P<0.001 |
| Acute kidney injury | 55 (28%) | 12 (9%) | 43 (66%) | 3 (11%) | P>0.05; P<0.001 |
| **Treatments** |  |  |  |  |  |
| Antiviral therapy | 187 (98%) | 123 (98%) | 64 (98%) | 13 (46%) | P<0.001; P>0.05 |
| Oxygen therapy | 182 (94%) | 118 (92%) | 64 (98%) | 28 (100%) | P>0.05; P>0.05 |
| Glucocorticoid therapy | 119 (62%) | 71 (55%) | 48 (74%) | 10 (36%) | P<0.05; P<0.05 |
| CRRT | 7 (4%) | 0 (0%) | 7 (11%) | 1 (4%) | P>0.05; P<0.001 |
| ECMO | 3 (2%) | 0 (0%) | 3 (5%) | 0 (0%) | P>0.05; P>0.05 |
| **Mechanical ventilation** |  |  |  |  |  |
| Non-invasive | 50 (26%) | 3 (2%) | 47 (72%) | 2 (7%) | P>0.05; P<0.001 |
| Invasive | 33 (17%) | 0 (0%) | 33 (51%) | 0 (0%) | P<0.05; P<0.001 |
| **Clinical outcomes** |  |  |  |  |  |
| Remained in hospital | 66 (34%) | 38 (30%) | 28 (43%) | 0 (0%) | P<0.001; P>0.05 |
| Discharged | 95 (49%) | 90 (70%) | 5 (8%) | 28 (100%) | P<0.001; P<0.001 |
| Death | 32 (17%) | 0 (0%) | 32 (49%) | 0 (0%) | P<0.05; P<0.001 |

Data are n (%). CP group=COVID-19 pneumonia group. OP group=other pneumonia group.

**Table 3:**
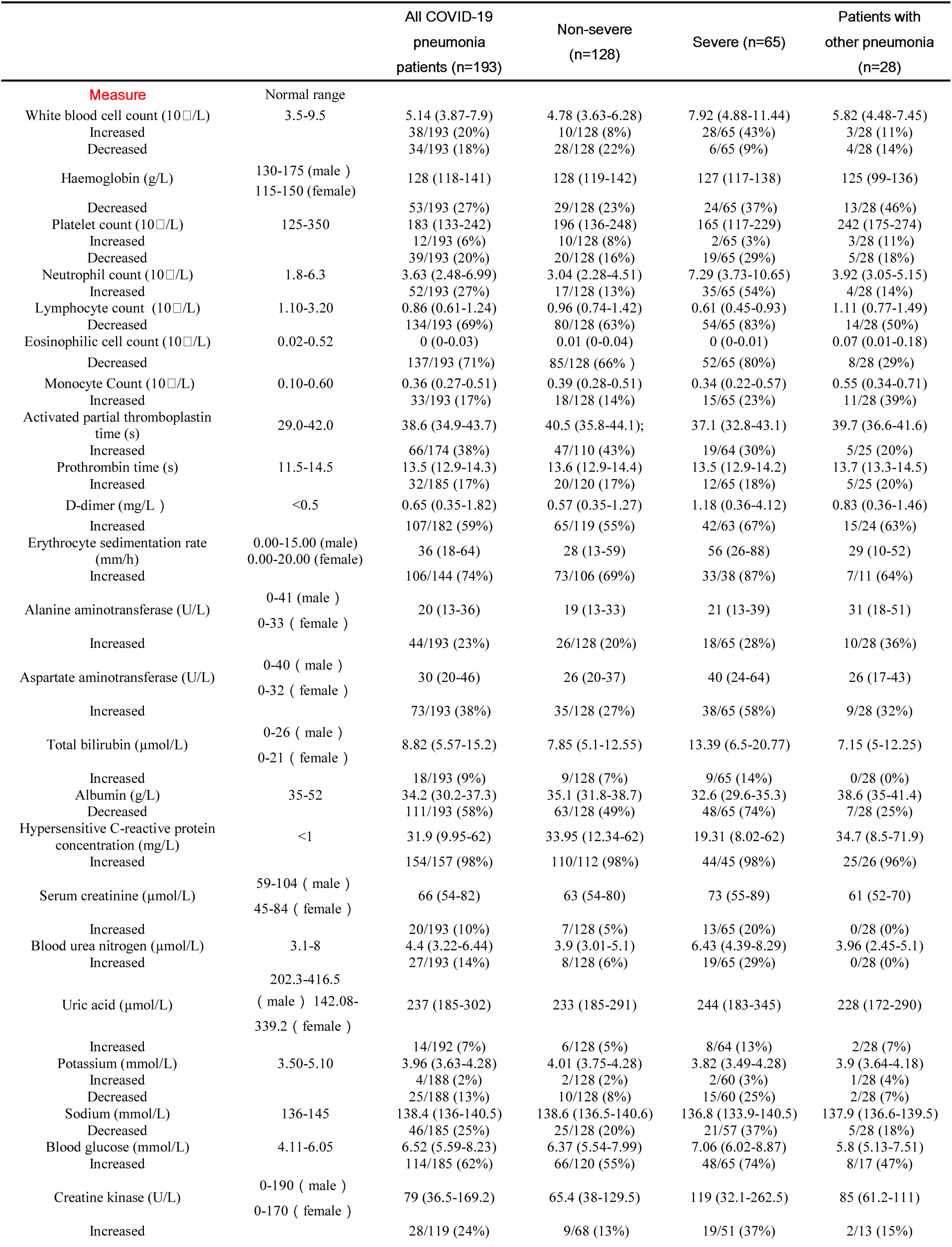
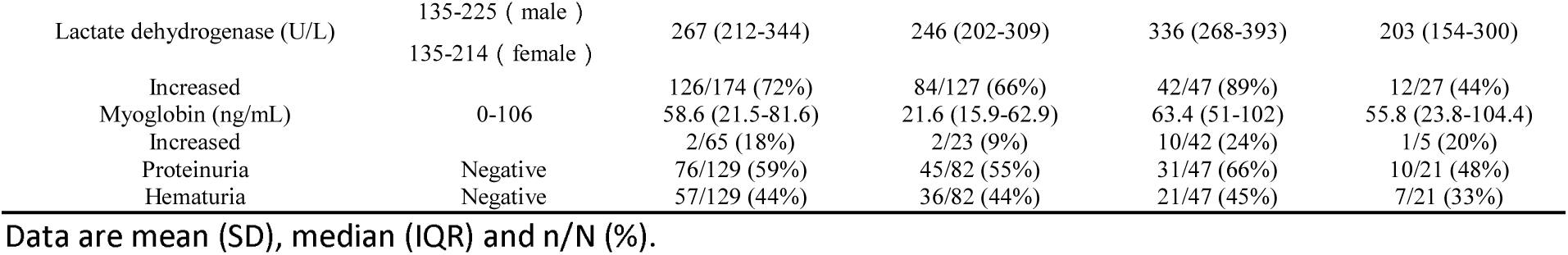
Laboratory findings of COVID-19 patients and other pneumonia patients on admission to hospital

According to the latest guidance provided by NHC China, following approaches were used to treat COVID-19 patients: (1) respiratory supports, including oxygen inhalation, mechanical ventilation and clean of the respiratory tract etc. (2) nutritional supports, including Albumin supplements etc. (3) drug therapies, including antiviral treatment, antibacterial treatment, antiasthmatic and expectorant treatment, glucocorticoid treatment etc. (4) other treatments of the potential serious complications (Table 2).

### Definition and interpretation

Stages of disease severity was determined according to the guidelines for diagnosis and treatment of COVID-19 published by NHC China on February 18, 2020 (6th Edition). Severe case was defined as either: (1) respiratory rate > 30/min, or (2) oxygen saturation ≤ 93%, or (3) PaO_2_/FiO_2_ ratio ≤ 300 mmHg. Lung imaging showed that the lesions progressed more than 50% within 24-48 hours, and these patients were also treated as severe cases. Critical severe case was defined as that including one criterion as follows: shock; respiratory failure requiring mechanical ventilation; combined with the other organ failure admission to intensive care unit (ICU).

According to the Kidney Disease: Improving Global Outcomes (KDIGO) criteria^9^, acute kidney injury (AKI) was defined as any of the following (not graded): (1) increase in serum creatinine (SCr) by ≥0.3 mg/dl (≥ 26.5 μmol/l) within 48 hours; or (2) increase in SCr to ≥ 1.5 times of baseline, which is known or presumed to have occurred within the prior 7 days; or (3) urine volume < 0.5 ml/kg/hour for 6 hours.

Acute respiratory distress syndrome (ARDS) was defined according to the Berlin definition^10^.

### CT examination and image acquisition

All CT data were collected from patients in Wuhan Tongji Hospital. The inclusion criteria were specified as: (1) kidneys were measurable within the scan range based on mediastinal window; (2) Patients has no known history ofkidney diseases. CT data were acquired by a SIEMENS scanner (SIEMENS/SOMATOM Definition AS+). Parameters were: rotation time, 0.5 seconds; detector pitch, 1.2; automatic tube current; tube voltage, 120 kV; slice thickness, 8 mm; reconstruction thickness, 1mm. No intravenous contrast enhancing agent was used. In addition to the conventional CT mean value analysis, CT texture analysis^11^ (CTTA) was also used for quantification. An abdominal radiologist, who was blinded to the study grouping, transferred all original CT images to a dedicated computer for kidney texture analysis (Fire Voxel, New York University, NY, USA).

### Statistical analysis

Continuous variables were expressed as median (Inter-Quartile Range, IQR) and compared with the Mann-Whitney U test. Categorical variables were presented as number (%) and compared with the Chi-square test. The survival probability of COVID-19 patients in hospital were calculated using the Kaplan-Meier method, and the significance and the hazard ratio of the risk variables were estimated by fitting a Cox proportional hazards model. A significance level was considered statistically significant for P < 0.05. The software of MATLAB (version 2018b, MathWorks) was used for statistical analysis.

## Results

193 COVID-19-infected patients, aged 26-95 years (median: 57; IQR: 46-67; Table 1), were included in this study. Epidemiologically, there were four main clusters of patients: 116 (60%) were local residents of Wuhan with no direct exposure to the Huanan seafood market, 8 (4%) had recently been to Wuhan or other cities in Hubei province, 31 (16%) had close contacts with COVID-19 patients, including 16 (8%) were health-care workers, and the remaining 38 (20%) without any reported history of exposure. There was no gender difference (95 [49%] men versus 98 [51%] women) (Table 1). 105 (54%) patients had chronic diseases, including nervous system diseases, cardiovascular and cerebrovascular diseases, respiratory system diseases, digestive system diseases, urinary system diseases, reproductive system diseases and endocrine system diseases (Table 1). The most common symptoms at the onset of illness were fever (171 [89%]), cough (134 [69%]), fatigue (75 [39%]), dyspnea (70 [36%]), anorexia (62 [32%]), and expectoration(59 [31%]); less common symptoms were diarrhoea (35 [18%]), myalgia (26 [14%]), headache (20 [10%]), sore throat (13 [7%]), chest pain (13 [7%]), dizziness (13 [7%]), nausea (11 [6%]), abdominal pain (8 [4%]), rhinorrhoea (5 [3%]), vomiting (5 [3%]), and low back pain (one [1%]; Table 1). Comparing to non-severe COVID-19 patients (n = 128 [66%]), severe COVID-19 patients (n = 65 [34%]) were significantly older (median age, 66 [IQR, 56-71] versus 55 [IQR, 43-66]), and more likely to have chronic comorbidities especially respiratory system diseases (17 [26%] versus 10 [8%]). Many patients presented with complications, including ARDS (54 [28%]), acute cardiac injury (24 [12%]), septic shock (35 [18%]) and AKI (55 [28%]). Compared with non-severe COVID-19 patients, severe COVID-19 patients were more likely to have these complications (Table 2).

On admission, white blood cell counts were above the normal range in 38 (20%) COVID-19 patients and below the normal range in 34 (18%) COVID-19 patients on admission. Hemoglobin, lymphocyte count and eosinophilic cell count were sharply reduced in some COVID-19 patients (53 [27%], 134 [69%] and 137 [71%] respectively). Neutrophil counts were markedly increased in 52 (27%) COVID-19 patients. Platelets were below the normal range in 39 (20%) COVID-19 patients and above the normal range in 12 (6%) COVID-19 patients. Monocyte count was elevated in 33 (17%) COVID-19 patients (Table 3). Prothrombin time and activated partial thromboplastin time were prolonged in 32 (17%) and 66 (38%) COVID-19 patients respectively. 107 of 182 patients tested for D-dimer had levels above the normal range. Erythrocyte sedimentation rate and hypersensitive C-reactive protein were increased in most COVID-19 patients (106 in 144 and 154 in 157 respectively). The levels of alanine aminotransferase (ALT) and aspartate aminotransferase (AST) were elevated in 44 (23%) and 73 (38%) COVID-19 patients respectively. The level of albumin was reduced in 111 (58%) COVID-19 patients. There were alterations in renal function indicators on admission: levels of blood urea nitrogen and serum creatinine were increased in 27 (14%) and 20 (10%) COVID-19 patients respectively, suggesting the presence of renal impairment already before or at the moment of admission. Levels of uric acid, creatine kinase and lactate dehydrogenase were also increased in some COVID-19 patients. 129 COVID-19 patients were tested for routine urine, urinary protein of 76 (59%) patients and hematuria of 57 (44%) patients were positive (see details in Table 3). Chest CT images of 193 (100%) COVID-19 patients showed pneumonia. 152 (79%) patients showed multiple mottling and ground-glass opacity. Severe COVID-19 patients were more likely to exhibit multiple mottling and ground-glass opacity on chest CT than non-severe COVID-19 patients (Table 1).

All COVID-19 patients were treated in isolation, and were administered with empirical antibiotic treatment. 187 (98%) patients were treated with antiviral therapy, 182 (94%) with oxygen therapy, 119 (62%) with glucocorticoid therapy, 7 (4%) with continuous renal replacement therapy (CRRT), and 3 (2%) with extracorporeal membrane oxygenation (ECMO). 50 (26%) patients received noninvasive mechanical ventilation and 33 (17%) patients received invasive mechanical ventilation. By the end of March 4^th^, 66 (34%) patients are still being treated in hospital, 95 (49%) patients discharged, and 32 (17%) patients died (Table 2).

We next focused on the kidney functions over hospitalization in the following results.

### 1. 88/147 (60%) patients exhibited proteinuria, and 71/147 (48%) exhibited hematuria

The tests of protein and blood in urine were collected from 147 patients (Fig. 1). Proteinuria was found in 88/147 (60%) patients and hematuria was found in 71/147 (48%) patients. For proteinuria, the semiquantitative result showed (±) in 31 patients (21%), (+) in 39 patients (27%), (++) in 15 patients (10%), and (+++) in 3 patients (2%) (Fig. 1B), with no significant difference among non-severe, severe (including death) cases and the patients with other pneumonia (Fig. 1A), suggesting the presence of kidney dysfunctions commonly in both non-severe and severe patients. Similarly, for hematuria, we found (±) in 21 patients (14%), (+) in 21 patients (14%), (++) in 16 patients (11%), and (+++) in 13 patients (9%), with severe patients being more significantly (P < 0.05) serious than non-severe ones. Note that, as mentioned above, the detected urine protein or blood was found in some patients (59% [76/129] for proteinuria and 44% [57/129] for hematuria) even on the first day of admission, offering a potential time window for starting interventions to protect kidney functions.

**Figure 1.**
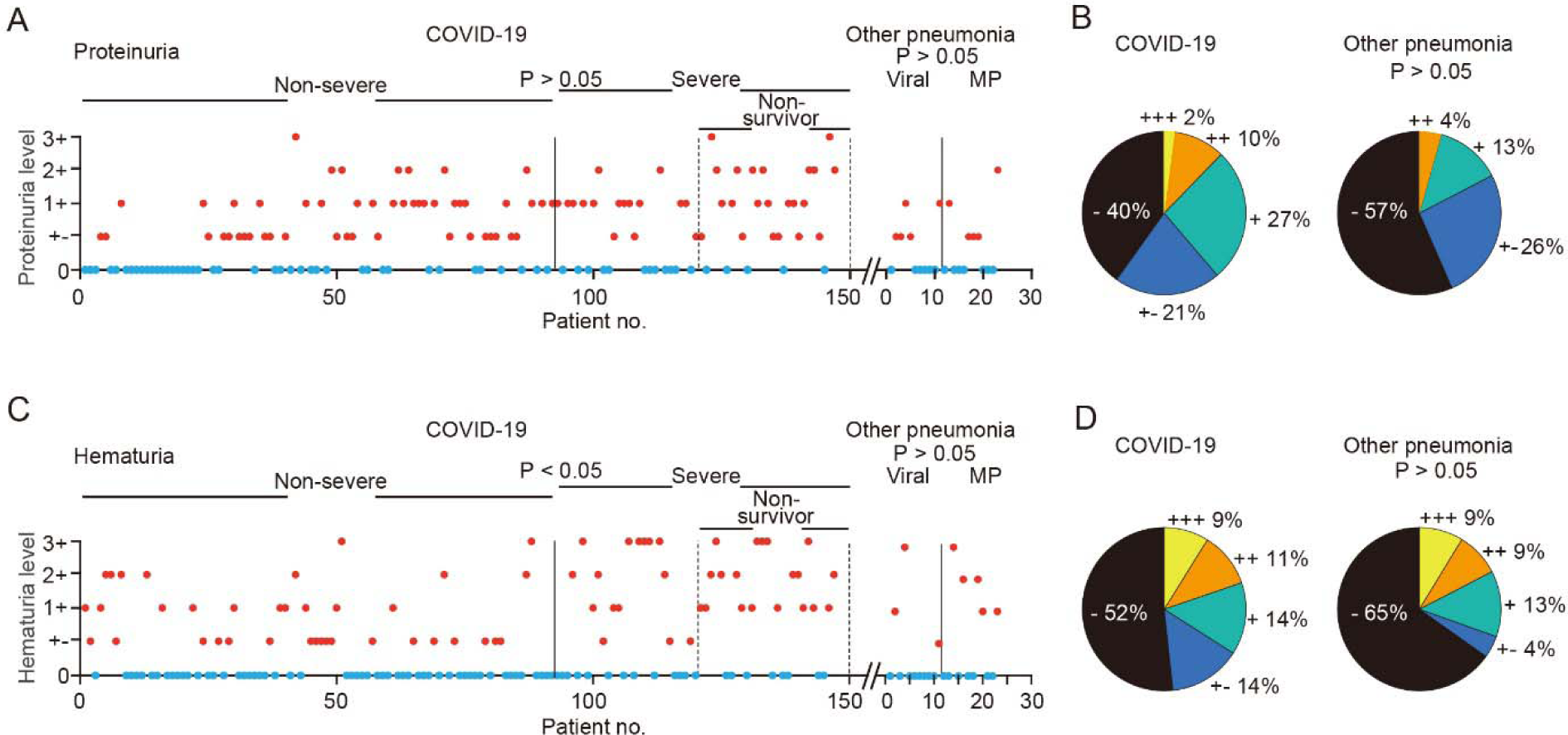
Proteinuria and hematuria in COVID-19 patients and other pneumonia patients. A, Levels for COVID-19 patients (n = 147) and other pneumonia patients (n = 23) tested for urine protein. Red circles correspond to proteinuria. The number of non-severe COVID-19 patients was 92. B, Percentages of all patients in the panel A. Comparison between the groups of, non-severe and severe: P > 0.05; COVID-19 and other pneumonia: P > 0.05, Chi-square test. Note that 0 represents negative. C, Levels for COVID-19 patients (n = 147) and other pneumonia patients (n = 23) tested for hematuria. Red circles correspond to hematuria. The number of non-severe COVID-19 patients was 92. D, Percentages of patients in the panel C. The number of severe COVID-19 patients was 55, including 27 non-survivors, and the other pneumonia patients included viral pneumonia patients (n = 11) and mycoplasma (MP) pneumonia patients (n = 12) in panel A and C. Comparison between the groups of, non-severe and severe: *P < 0.05, COVID-19 and other pneumonia: P > 0.05, Chi-square test. Note that 0 represents negative.

### 2. Blood urea nitrogen (BUN): 59/193 (31%) patients exhibited an elevated level of BUN

In COVID-19 patients with increased BUN levels (59/193 cases, 31%), the peak BUN levels ranged from −0.45 to 7.40 of the normalized value, with a median value of 0.51 of the normalized value (normal value before normalization: 3.1-8 mmol/L; Fig. 2A, B). In contrast, only one (4%) of other pneumonia patients exhibited an elevated level of BUN (Fig. 2B). In 169 COVID-19 cases and 17 other pneumonia cases, we were able to collect the data for the change in BUN levels over days (Fig. 2C), and this result showed that the duration from the onset of admission to the presence of the BUN increase was in the range of 0 to 16 days (median 2 days). Note that severe patients (including deceased cases) had a significantly higher level of BUN than that of non-severe cases or of other pneumonia ones (p < 0.001). In particularly, the high levels of BUN were common in the severe and deceased cases across the course of disease.

**Figure 2.**
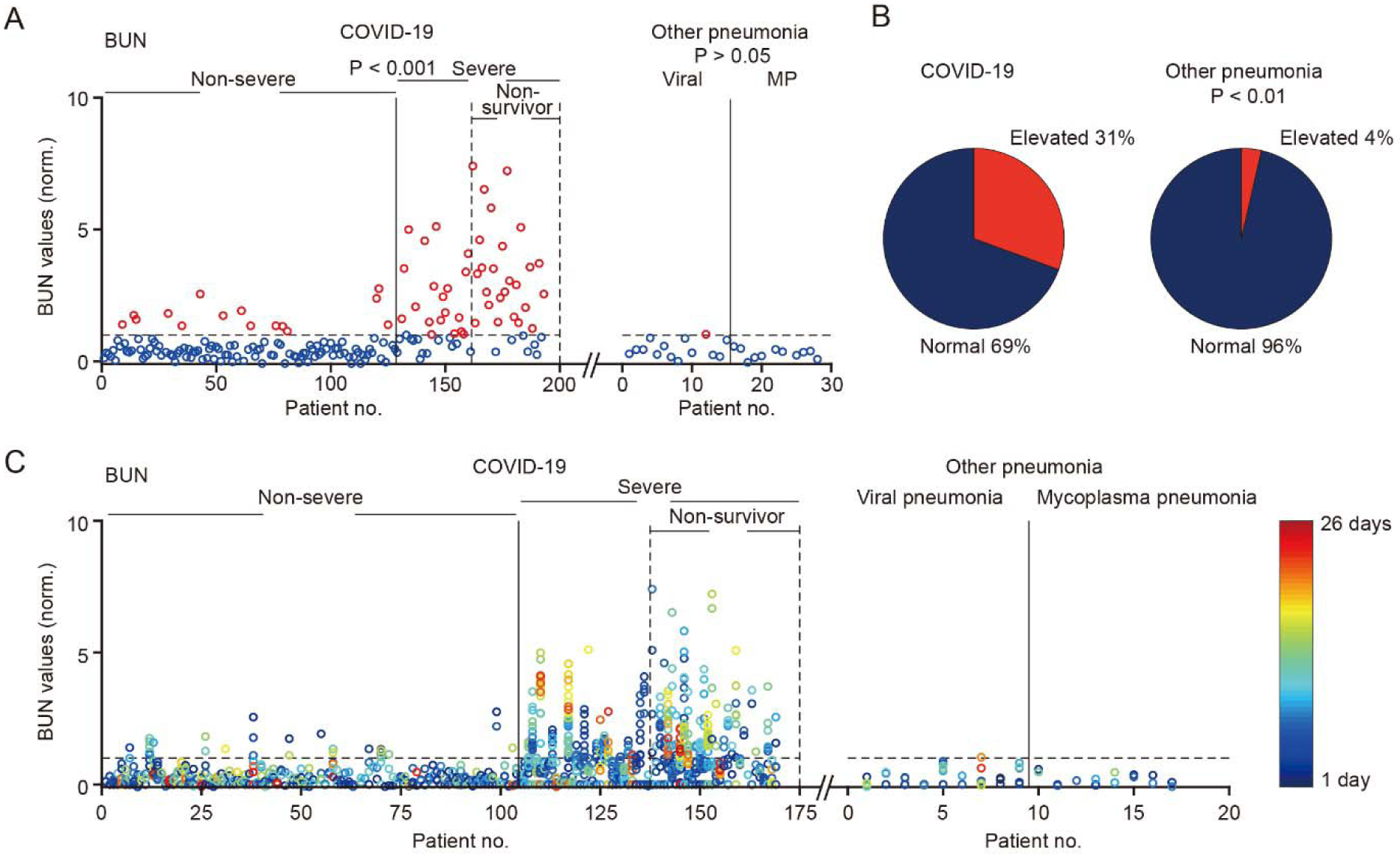
The levels of blood urea nitrogen (BUN) in in COVID-19 patients and other pneumonia patients. A, Values for COVID-19 patients (n = 193) and other pneumonia patients (n = 28) tested for BUN. Red circles correspond to elevated levels of BUN. The number of non-severe COVID-19 patients was 128. The number of severe COVID-19 patients was 65, including 32 non-survivors. The other pneumonia patients included viral pneumonia patients (n = 15) and mycoplasma (MP) pneumonia patients (n = 13). Comparison between the groups of, non-severe and severe: ***P < 0.001, non-severe and other pneumonia: P > 0.05, Wilcoxon rank-sum test. B, Percentages of all patients in the panel A exhibited normal or elevated BUN. C, BUN values for COVID-19 patients (n = 169) and other pneumonia patients (n = 17) tested over multiple days (> 1 day). The color-coded circles correspond to the BUN values of different days. The number of non-severe COVID-19 patients was 104. The number of severe COVID-19 patients was 65, including 32 non-survivors. The other pneumonia patients (n = 17) included 9 viral pneumonia patients and 9 mycoplasma pneumonia patients. Note that the dotted lines indicate the normal level that has been normalized to 1.

### 3. Plasma creatinine (SCr): 43/193 (22%) patients exhibited an elevated level of SCr

In COVID-19 patients with increased SCr levels (43/193 cases, 22%), we found the peak SCr levels ranged from - 0.49 to 7.59 of the normalized value (median 0.49; normal value before normalization: 59-104 μmol/L for male, 45-84 μmol/L for female) (Fig. 3A, B). In 169 COVID-19 cases and 17 other pneumonia cases, we were able to monitor the change in SCr levels over multiple days (Fig. 3C). We found that the duration from the onset of admission to the presence of the SCr increase was in the range of 0 to 20 days (median 5 days). Note that, similar to the level of BUN, severe patients (including deceased ones) had a significantly higher level of SCr than that of non-severe cases or of other pneumonia ones (P < 0.001). Importantly, the high levels of SCr were commonly present in the severe cases across the course of disease, supporting the SCr level as a risk factor predicting mortality in coronavirus-infected patients^8^. In contrast, for other pneumonia cases, there was no one with an elevated level of SCr.

**Figure 3.**
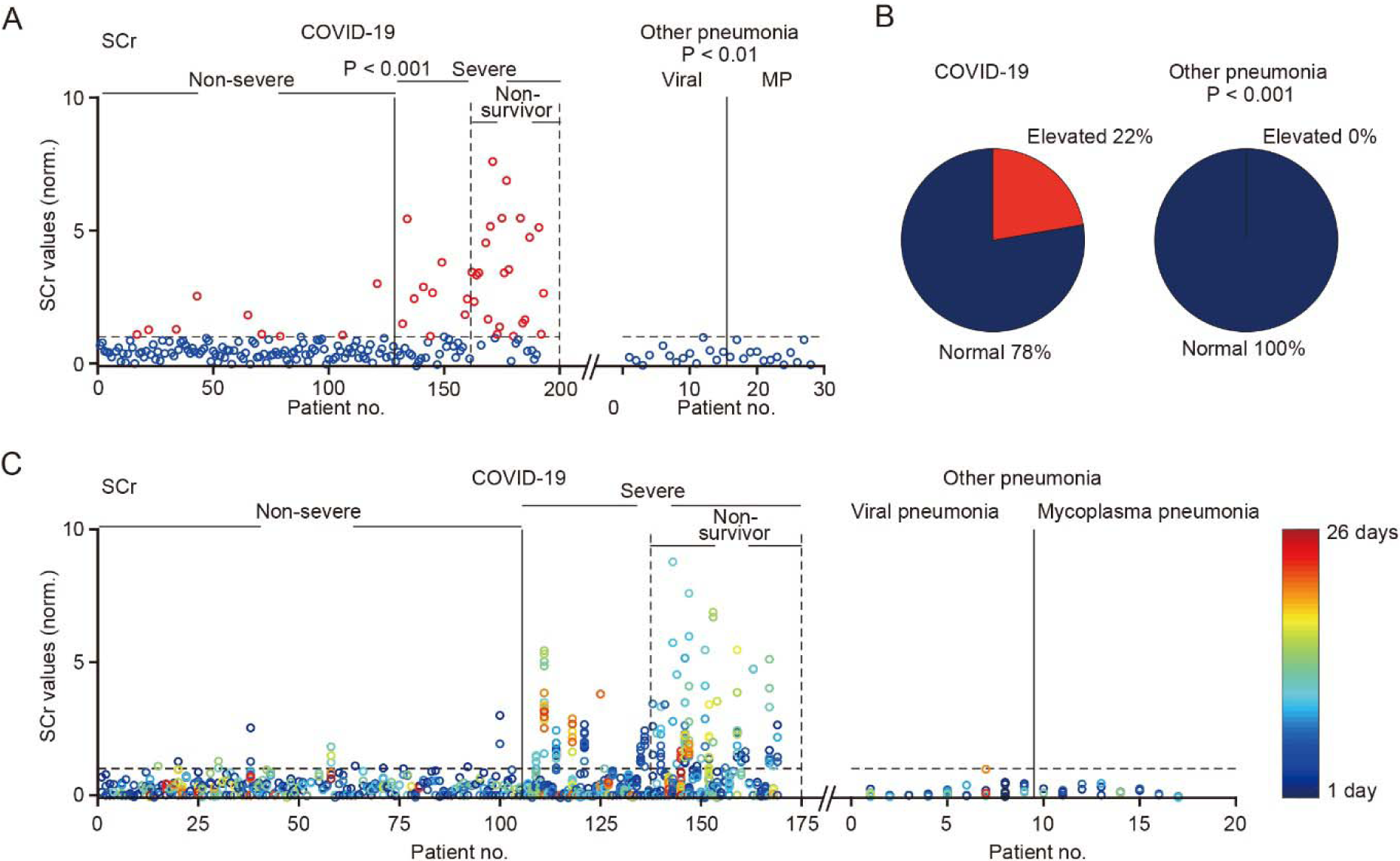
The levels of serum creatinine (SCr) in in COVID-19 patients and other pneumonia patients. A, Values for COVID-19 patients (n = 193) and other pneumonia patients (n = 28) tested for SCr. Red circles correspond to elevated levels of SCr. The number of non-severe COVID-19 patients was 128. The number of severe COVID-19 patients was 65, including 32 non-survivors. The other pneumonia patients included viral 15 pneumonia patients (n = 15) and 13 mycoplasma (MP) pneumonia patients. Comparison between the groups of, non-severe and severe: ***P < 0.001, non-severe and other pneumonia: **P < 0.01, Wilcoxon rank-sum test. B, Percentages of all patients in the panel A exhibited normal or elevated SCr. C, SCr values for COVID-19 patients (n = 169) and other pneumonia patients (n = 17) tested over multiple days. The color-coded circles correspond to the SCr values of different days. The number of non-severe COVID-19 patients was 104. The number of severe COVID-19 patients was 65, including 32 non-survivors. The other pneumonia patients included viral pneumonia patients (n = 9) and mycoplasma pneumonia patients (n = 8). Note that the dotted lines indicate the normal level that has been normalized to 1.

### 4. 39/192 (20%) patients exhibited an elevated level of uric acid (UA) and 128/182 (70%) exhibited an elevated level of D-dimer (DD)

We found 20% (39/192) COVID-19 patients with increased UA levels and 70% (128/182) with increased DD levels. The peak UA levels ranged from −0.54 to 4.59 of the normal value, with a median of 0.51 of the normal value (normal value before normalization: 202.3-416.5 μmol/L for male, 142.08-339.2 μmol/L for female; Fig. 4A, B), and the peak levels for DD was from 0.04 to 200.6 of the normal level, with a median of 2.52 (normal value before normalization: 0-0.5 mg/L; Fig. 5A, B). In contrast, only 11% of other pneumonia patients (n = 28) exhibited an elevated level of UA (Fig. 4B), while these other pneumonia cases (n = 24) commonly exhibited elevated levels of DD (63% of cases; Fig. 5B). In some cases, we were able to collect the data for the change in UA or DD levels over days (Fig. 4C and 5C), and this result showed that the duration from the onset of admission to the presence of the UA or DD increase was in the range of 0 to 20 days (median 7 days) or 0 to 22 days (median 0 days). Note that severe patients (including deceased cases) had a significantly higher level of UA or DD than that of non-severe cases (UA: P < 0.001; DD: P < 0.001) or of other pneumonia ones (UA: P < 0.01; DD:P < 0.001). In particularly, the high levels of DD were common in the severe and deceased cases across the course of disease, consistent with recent reports that a high level of DD could be an important mortality risk factor^5,12^.

**Figure 4.**
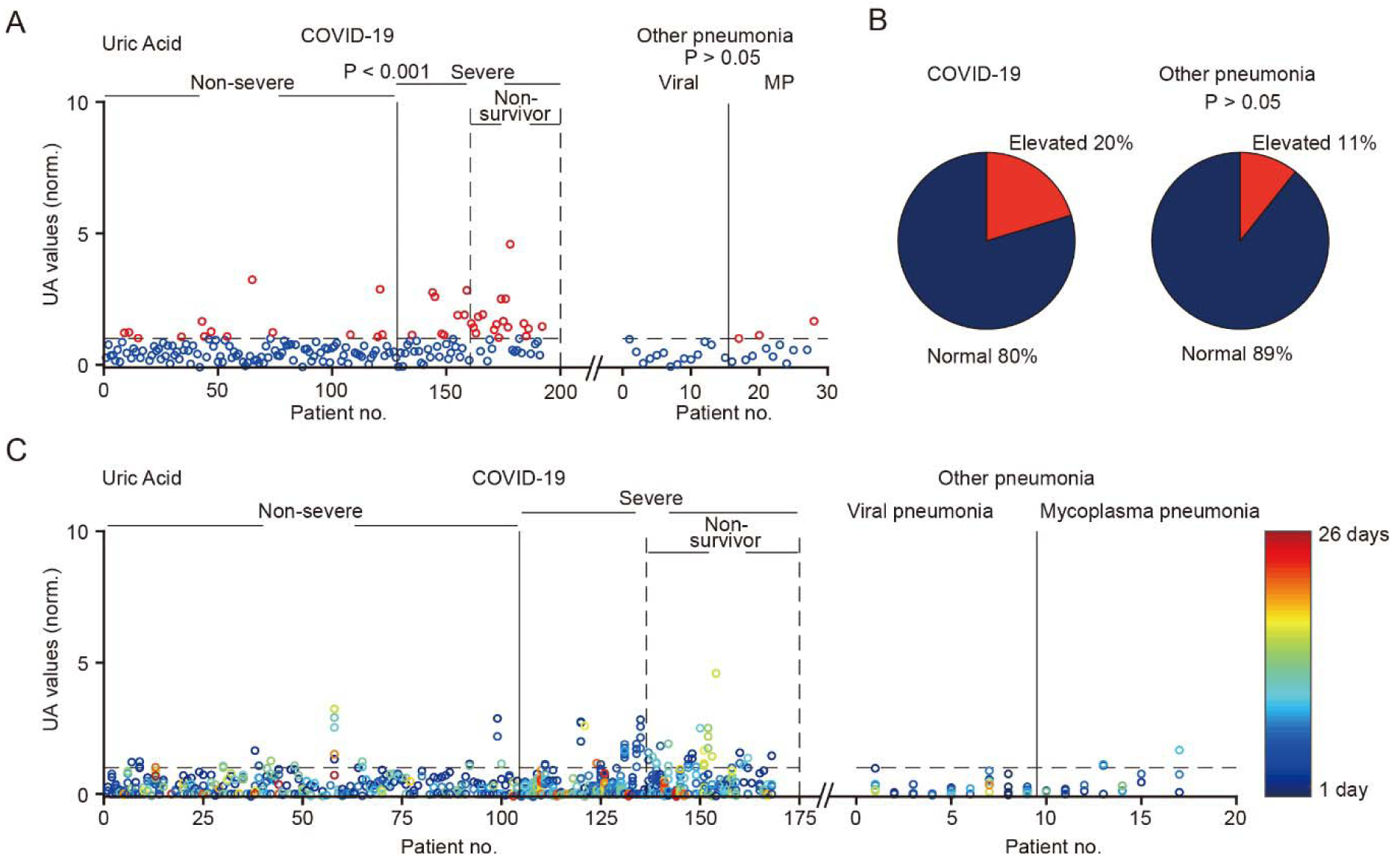
The levels of uric acid (UA) in in COVID-19 patients and other pneumonia patients. A, Values for COVID-19 patients (n = 192) and other pneumonia patients (n = 28) tested for UA. Red circles correspond to elevated levels of UA. The number of non-severe COVID-19 patients was 128. The number of severe COVID-19 patients was 64, including 32 non-survivors. The other pneumonia patients included viral pneumonia patients (n = 15) and mycoplasma (MP) pneumonia patients (n = 13). Comparison between the groups of, non-severe and severe: ***P < 0.001; non-severe and other pneumonia: P > 0.05, Wilcoxon rank-sum test. B, Percentages of all patients in the panel A exhibited normal or elevated UA. C, UA values for COVID-19 patients (n = 168) and other pneumonia patients (n = 17) tested over multiple days. The color-coded circles correspond to the UA values of different days. The number of non-severe COVID-19 patients was 104. The number of severe COVID-19 patients was 64, including 32 non-survivors. The other pneumonia patients were viral pneumonia patients (n = 9) and mycoplasma pneumonia patients (n = 8). Note that the dotted lines indicate the normal level that has been normalized to 1.

**Figure 5.**
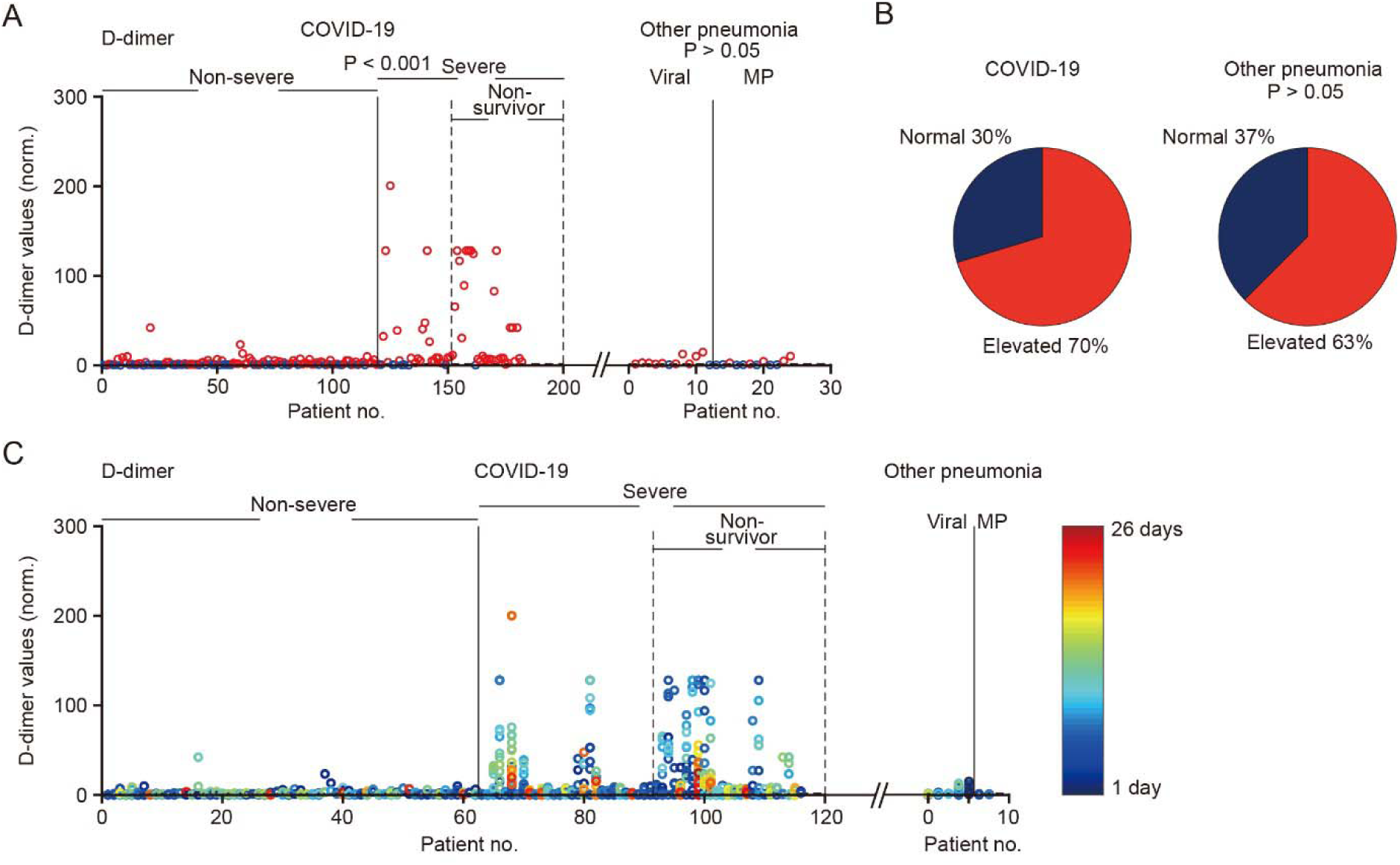
The levels of D-dimer in in COVID-19 patients and other pneumonia patients. A, Values for COVID-19 patients (n = 182) and other pneumonia patients (n = 24) tested for D-dimer. Red circles correspond to elevated levels of D-dimer. The number of non-severe COVID-19 patients was 119. The number of severe COVID-19 patients was 32, including 31 non-survivors. The other pneumonia patients were viral pneumonia patients (n = 12) and mycoplasma (MP) pneumonia patients (n = 12). Comparison between the groups of, non-severe and severe: ***P < 0.001, non-severe and other pneumonia: P > 0.05, Wilcoxon rank-sum test. B, Percentages of all patients in the panel A exhibited normal or elevated D-dimer. C, UA values for COVID-19 patients (n = 116) and other pneumonia patients (n = 7) tested over multiple days. The color-coded circles correspond to the D-dimer values of different days. The number of non-severe COVID-19 patients was 62. The number of severe COVID-19 patients was 54, including 25 non-survivors. The other pneumonia patients were viral pneumonia patients (n = 5) and mycoplasma pneumonia patients (n = 2). Note that the dotted lines indicate the normal level that has been normalized to 1.

### 5. CT data: 106/110 patients exhibited radiographic abnormalities of kidney

In 110 COVID-19 patients and 24 patients with other pneumonia from Wuhan Tongji hospital, we were able to collect the plain CT scan results of the parenchyma of kidney in the same chest CT scan datasets because the scanned volume included a part of kidney. The mean CT value and CT texture analysis (CTTA) parameters (skewness, kurtosis and entropy) of the renal parenchyma were measured on the image of the largest layer of thekidney^13,14^. We found that the mean CT value of COVID-19 patients was in the range of 17.0-36.0 HU, with amedian of 27.3 HU (see the example of two patients in Figure 5A and B), which was significantly lower than the healthy control group who had no kidney diseases (n = 109; 33.2 HU) in Wuhan Tongji Hospital. This mean CT value was also lower than that of normal kidney CT value from a previous study (38.0 HU)^15^. 106/110 patients had a smaller value than 33.2 HU (Fig. 5C). In addition, the mean CT value of COVID-19 patients was also lower than that of patients with other pneumonia (32.8 HU). The skewness value (−0.11 [−0.26 to −0.02]) and entropy value (3.9 [3.8 to 4.0]) of COVID-19 patients were lower than the healthy control group (Skewness (−0.04 [−0.13 to 0]); entropy (3.9 [3.8-4.0])), while the kurtosis value of COVID-19 patients (0.33 [0.16 to 1.02]) was higher than the healthy control group (0.17 [0.03 to 0.47]). Principal component analysis (PCA) was used for dimensionality reduction of the bilateral renal mean CT value and CTTA parameters. The first two components explained 99.0% of all variability. The bilateral renal mean CT value and CTTA parameters of COVID-19 patients were significant different from those of healthy controls or patients with other pneumonia (Figure 6G; p < 0.001, two-way ANOVA). Together, these results indicate that inflammation and edema of the renal parenchyma may commonly occur in COVID-19 patients.

**Figure 6.**
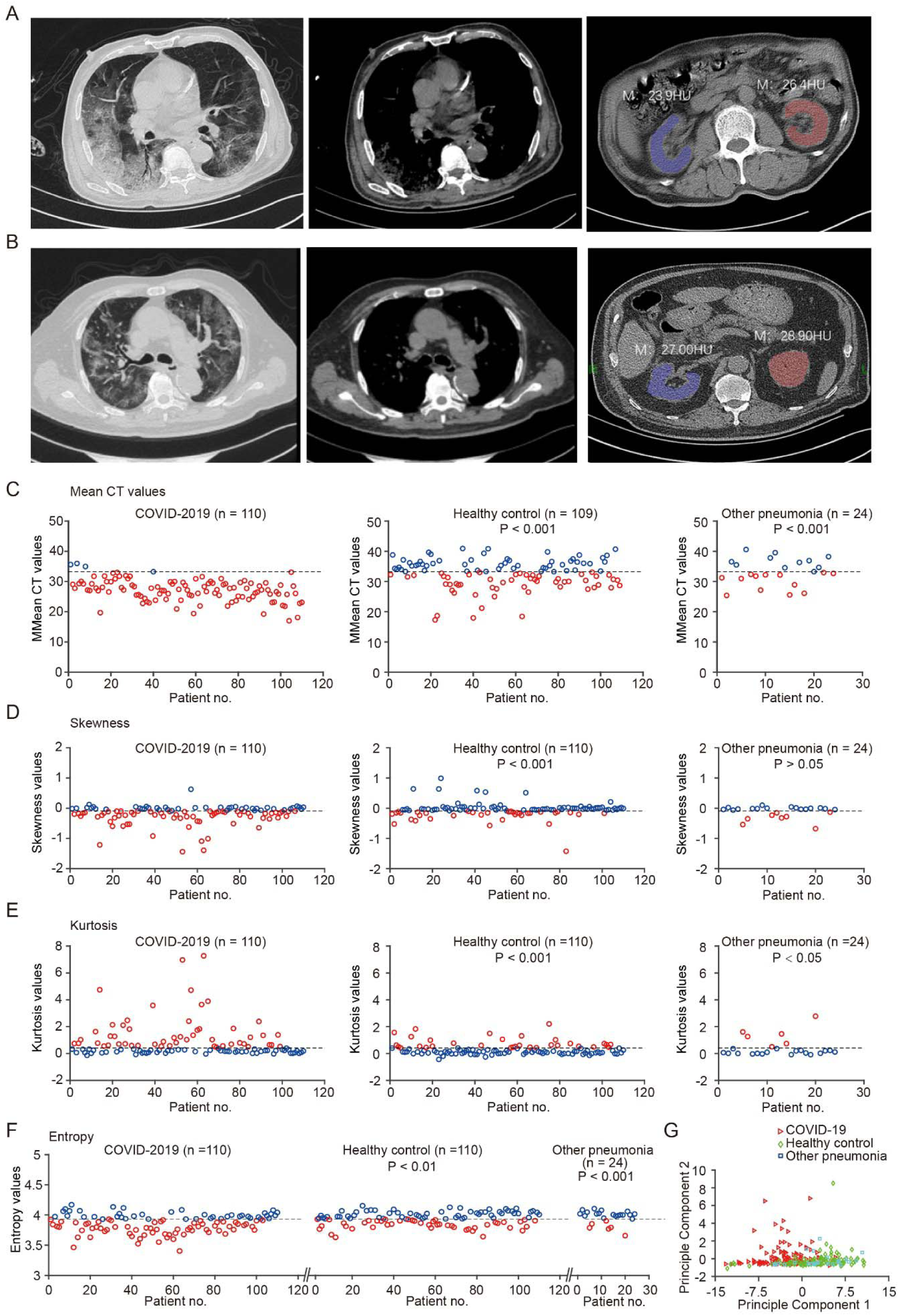
Radiographic abnormalities of the kidneys by CT scan. A and B, CT examples of two different patients (A: 71-year old male, B: 70-year old male), respectively. Left, the largest layers of the lung show diffuse bilateral confluent and patchy ground-glass. Middle, the largest layers of the mediastinum. Right, column, the largest layers of kidney and CT values of bilateral kidney. C, Mean CT values of bilateral kidney. Red circles correspond to the decreased levels (Comparison between COVID-19 patients and health control subjects, P < 0.001; Comparison between COVID-19 patients and other pneumonia patients, P < 0.001). D, Skewness values of bilateral kidney. Red circles correspond to the decreased levels (Comparison between COVID-19 patients and health control subjects, P < 0.001; Comparison between COVID-19 patients and other pneumonia patients, P > 0.05). E, Kurtosis values of bilateral kidney. Red circles correspond to the elevated levels (Comparison between COVID-19 patients and health control subjects, P < 0.001; Comparison between COVID-19 patients and other pneumonia patients, P < 0.05). F, Entropy values of bilateral kidney. Red circles correspond to the decreased levels (Comparison between COVID-19 patients and health control subjects, P < 0.01; Comparison between COVID-19 patients and other pneumonia patients, P < 0.001). G, Principal component analysis (PCA) shows that bilateral renal CT texture analysis (CTTA) parameters of COVID-19 patients were significantly different from those of either healthy controls or patients with other pneumonia (Fig. 6G, P < 0.001, two-way ANOVA). Note that the dotted lines indicate the normal levels of healthy control individuals.

### 6. Kidney functions associated with survival probability of COVID-19 patients

We next performed a survival analysis for COVID-19 patients regarding the association of death probability with multiple kidney function indicators, including proteinuria, hematuria, BUN, SCr, UA, D-dimer (Figure 7A-F). We adopted univariate Cox regression analyses and showed that death of COVID-19 patients was significantly associated with the elevated levels of proteinuria (P < 0.05), hematuria (P < 0.01), BUN (P < 0.001), SCr (P < 0.001), UA (P < 0.001) and D-dimer (P < 0.05) respectively. Moreover, the estimated hazard ratio indicates that the mortality risk for COVID-19 patients with AKI was significantly higher than (∼5.3 times, P < 0.001) the mortality risk for patients without AKI. In contrast, the mortality risk for COVID-19 patients with chronic illnesses was not significantly higher than (∼1.5 times, p > 0.05) the mortality risk for patients without comorbidity.

**Figure 7.**
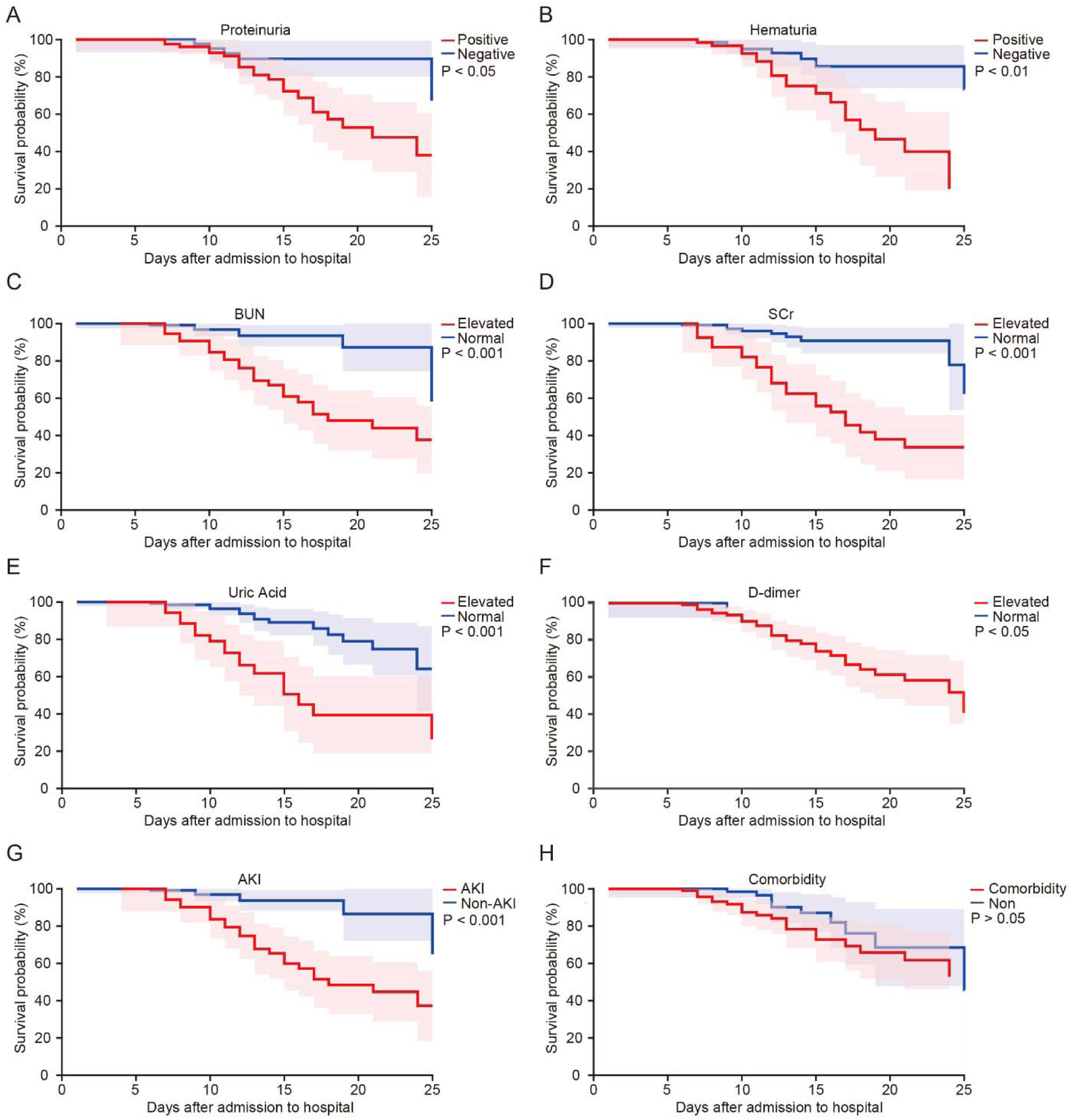
Survival probability for the subgroups of the COVID-19 patients with different kidney dysfunction indicators, proteinuria, hematuria, BUN, SCr, UA, D-dimer, acute kidney injury (AKI) AND comorbidity (Univariate Cox regression analysis).

## Discussion

Although the lung was commonly accepted as the primary target organ of SARS-CoV-2 infection as demonstrated by the early autopsy reports^16^, we found that COVID-19 patients also generally exhibited kidney dysfunctions as characterized by the presence of proteinuria & hematuria (Figure 1) and also by the elevated levels of BUN (Figure 2), SCr (Figure 3), UA (Figure 4) and DD (Figure 5). In particular, BUN and SCr (the relevant indicator of AKI^8^) levels were higher in non-severe COVID-19 patients than in patients of other commonly known pneumonia, albeit the respiratory symptoms were largely indifferent at this stage. This result was further supported by the analysis of kidney CT images (to our knowledge, this is the first report of kidney CT data in COVID-19 patients^17^; Figure 6) that the group of non-severe COVID-19 patients were significantly distinguishable from either the group of other pneumonia or the healthy control group. These results together showed the general presence of kidney dysfunctions in COVID-19 patients, which for most non-severe patients were mild and not diagnosed as AKI (Table 1, 9%, 12 of 128 with AKI). In contrast, AKI was found in a significant fraction of severe patients (Table 1, 66%, 43 of 65 with AKI). In addition, the factors for evaluating kidney functions including BUN, SCr, UA and DD had significantly higher levels in severe patients including deceased cases than those in non-severe ones. And all these factors were significantly associated with the death of COVID-19 patients. Importantly, we should mention that a high level of DD as a potential risk factor has been also reported in COVID-19 patients very recently^5^. In addition, our survival analysis showed a striking contrast that COVID-19 patients with AKI had ∼ 5.3 times mortality risk higher than those without AKI (Figure 7G). In contrast, patients with the above-mentioned chronic illnesses had only on average ∼ 1.5 times mortality risk (Figure 7H). Therefore, the presence of kidney dysfunctions in COVID-19 patients is an important negative prognostic factor for survival.

The occurrence of kidney dysfunctions in COVID-19 patients might be explained by the kidney-lung crosstalk theory^18-21^ because of the following reasons. First, the SARS-CoV-2 uses ACE2 (angiotensin-converting enzyme 2) as a cell entry receptor^6^, which was not exclusively expressed in the respiratory organs in humans but also in the kidney with a much higher level than that in the lung. Thus, it is possible that SARS-CoV-2 could also attack renal tubular epithelial cells in addition to attacking lung epithelial cells^22^. Consistent with this possibility, a recent preprinted report analyzed the histology of renal tissues from autopsies and found acute renal tubular damage in six COVID-19 cases^23^. Second, kidney dysfunctions could accelerate the inflammation progress started at the lung, not just as a collateral damage of lung-derived inflammation. On the one hand, inflammatory reactions following lung impairments could damage the kidney; on the other hand, the injury and death of renal tubular epithelial cells could also cause severe damage of the lung and other organs through a large amount of inflammatory substances^19,24^. Over a certain critical point, the kidney-lung crosstalk could lead to an irreversible self-amplifying cytokine storm that rapidly induces multi-organ failure and death.

The limitations of our study and implications are as follows. First, clinical data regarding kidney functions were incomplete or absent in a substantial fraction of COVID-19 patients in the overburdened local medical system during the outbreak, thus we only included patients with complete data on kidney functions (except for CT data) in this study. The sampling bias was not subjective, because the existence and collection of our data was irrelevant to clinical considerations on whether COVID-19 patients had kidney diseases or not. In particular, kidney CT data were collected not from kidney-specific scans but from the abundant image volume of chest scans. Nevertheless, our results, though largely comparable with others, should not be interpreted in the standard framework of epidemiology. This is because all our included hospitals were designated to preferentially treat severe COVID-19 patients while most non-severe patients were treated in temporarily-built field hospitals during the outbreak.Second, various drugs and interventions had been applied to COVID-19 patients, the use of which could have side-effects on kidney functions during the course of treatment, e.g., invasive mechanical ventilation^25^. However, regardless of what could have damaged the kidney and caused AKI in COVID-19 patients, our suggestions on exercising a high caution in monitoring kidney functions still stood. Third, this is a retrospective study that does not provide sufficient data regarding the effect of the suggested intervention method. In the meanwhile, our early alert on the kidney dysfunctions of COVID-19 patients and suggested interventions in the first preprinted version of this work^26^ have been already integrated into the official therapeutic guidelines announced by NHC China^27^ and practiced in more clinical cases.

Taken together, we suggest exercising a high degree of caution in monitoring kidney functions of COVID-19 patients regardless of comorbid chronic illnesses. To reduce the mortality risk for severely ill COVID-19 patients worldwide, we recommend to apply any supportive interventions that are potentially protective for kidney functions at the early stage of the disease, and importantly, to apply interventions including but not limited to e.g.,blood filtering and purification treatments^26^, and the use of any potential inhibitors against the adverse effects ofinflammatory cytokines^28,29^, as have been illustrated in details in the COVID-19 treatment guidelines published by NHC China^27^.

## Contributors

JuY, XWC, HJ, YiZ and MHa had the idea for and designed the study and had full access to all of the data in the study and take responsibility for the integrity of the data and the accuracy of the data analysis. JuY, XWC, HJ, YiZ and MHa drafted the paper. JuY, XWC, HJ, YiZ, JiY and XL did the analysis, and all authors critically revised the manuscript for important intellectual content and gave final approval for the version to be published. ZL, MW, JG, SS, JL, GD, YuZ, XW, ZZ, TW, MHu, XXC, YF, CL, HD, CX and YH collected the data and contributed to datainterpretation. All authors agree to be accountable for all aspects of the work in ensuring that questions related to the accuracy or integrity of any part of the work are appropriately investigated and resolved.

## Data Availability

Please find all data at the following link (code: 7hjn):

https://pan.baidu.com/s/1yDg_jwSv1RckHuVa6_dhYA

## Data sharing

The data of our study will be made available to others on reasonable requests to the corresponding author after publication. A proposal with detailed description of study objectives and statistical analysis plan will be needed for evaluation of the reasonability of requests. Additional materials might also be required during the process of evaluation.

## Acknowledgments

We thank all patients involved in this study. This study was supported by grants from the “1000-Talents Program for Young Scholars” of China (X. Chen) and the “100-Talents Program for Elite Engineers” of the CAS (H. Jia). X. Chen is a junior fellow of the CAS Center for Excellence in Brain Science and Intelligence Technology.

